# Prevalence of indications of alcohol and drug use among patients treated for injurious falls by Emergency Medical Services

**DOI:** 10.1101/2024.06.03.24308063

**Authors:** Nicole G. Itzkowitz, Kathryn G. Burford, Remle P. Crowe, Henry E. Wang, Alexander X. Lo, Andrew G. Rundle

## Abstract

**Objective:** To describe the distribution of alcohol and drug involvement in injurious falls by location and subtype of fall.

**Methods:** Using the 2019 National Emergency Medical Services Information System (NEMSIS) dataset we identified 1,854,909 patients injured from falls requiring an Emergency Medical Services (EMS) response and determined the fall location (e.g. indoors or on street/sidewalk) and the EMS clinician’s notation of alcohol or drug involvement. We analyzed substance involvement by fall subtype, location of fall and patient demographics.

**Results:** Overall, for 7.4% of injurious falls there was a notation of substance use: 6.5% for alcohol alone, 0.6% for drugs and 0.3% for alcohol and drugs. 21.2% of falls that occurred on a street or sidewalk had a notation of substance use; alcohol use alone for 18.5% of falls, drugs alone for 1.7% of falls and alcohol and drugs for 0.9% of falls. Substance use prevalence was highest, at 30.3%, in the age group 21 to 64 years, for falls occurring on streets and sidewalks, without syncope or heat illness as contributing factors; alcohol use alone for 26.3%, drugs alone for 2.6%, and alcohol and drugs for 1.4%. Reported substance use involvement was more frequent for men compared to women for each location type.

**Conclusions:** Overall, 1-in-5 injurious falls on streets and sidewalks and requiring EMS attention involved substance use, and these numbers likely underestimate the true burden. As cities seek to expand nightlife districts, design strategies to protect pedestrians from falls should be enacted.

## Introduction

The association between alcohol consumption and increased injuries from falls is well established. (1-5) A recent systematic review of substance use among falls patients recommended that research address the prevalence of substance use by fall type and the prevalence of drug use as well as alcohol use. (6) Prior studies have rarely distinguished between falls that occur in the home and falls that occur outdoors, and particularly those occurring on streets and sidewalks. (7) This distinction is important because current fall prevention guidelines focus on indoor falls and not on falls occurring outdoors. (8-11) However, there are policy tools, such as stricter enforcement of existing licensing laws, that can be used to influence the extent of alcohol consumption in public spaces, bars and restaurants and that can be used to improve outdoor pedestrian safety environments around bars and restaurants and in nightlife districts. (12) It has been estimated that over 500,000 falls requiring at least some form of medical attention occur on streets and sidewalks annually. (13) To our knowledge national or regional estimates of the number of alcohol and/or drug involved injurious falls that occur on streets and sidewalks do not exist. However, data on the number of pedestrians under the influence of alcohol who are injured or killed by motor vehicles suggest that pedestrians under the influence of alcohol constitute a significant portion of vulnerable road users. (14-16) The National Highway Traffic Safety Administration (NHTSA) reported that 31% of pedestrians who were killed in a traffic crash had a BAC of 0.08 g/dL or higher in 2019. As such, it is likely that a large number of injurious falls occurring outdoors involve pedestrians who are alcohol impaired.

The only extant public health messaging on substance use by pedestrians comes from the NHTSA which simply advises pedestrians to “avoid alcohol and drugs when walking; they impair your judgment and coordination”. (16, 17) The most recent set of policy and design recommendations for reducing injuries among intoxicated pedestrians was a 1996 report from Monash University. (18) This is a concern because many cities, as part of economic development plans to revitalize urban centers, are now developing and promoting nightlife and nightlife districts, prominently featuring alcohol-serving establishments, which encourage patrons to walk between nearby alcohol serving venues. (19-22) In addition, nightlife districts and the nighttime economy are also associated with illicit drug use. (23-25) Furthermore, multiple jurisdictions have open container laws, that allow drinking on streets and sidewalks, and to-go alcohol purchases, policies that also encourage walking and drinking. (20, 26, 27) Additionally, multiple states have legalized or decriminalized cannabis and within these states retail outlets selling cannabis have proliferated, with some allowing on-site consumption, and use of cannabis in public spaces is becoming common. (28-31) These economic and policy initiatives are likely to increase the number of pedestrians under the influence of alcohol and at risk of falls and injuries from motor vehicles. (12, 20, 32, 33)

To support advances in polices and urban design strategies to prevent pedestrian falls, there is a need for data describing the burden of alcohol and drug-related injurious falls (6). As an initial effort to describe this burden, we used the 2019 National Emergency Medical Services Information System (NEMSIS) public-use research dataset to assess the prevalence of indicators for substance use among patients treated for injurious falls by Emergency Medical Services (EMS).

## Methods

### Study design and setting

This cross-sectional study of EMS records from the 2019 National Emergency Medical Services Information System (NEMSIS) Public-Release Research Dataset included 1,854,909 occurrences of falls requiring EMS response across US states and territories. (7)

### Data source

NEMSIS is the national system to collect and standardize data from EMS agencies across the US.

The National Highway Traffic Safety Administration Office for Emergency Medical Services provides the NEMSIS data as a public use, de-identified, Health Insurance Portability and Accountability Act exempt dataset hosted by the University of Utah, therefore further institutional review board (IRB) review was not requested. (https://nemsis.org/using-ems-data/). (34-36) The use of NEMSIS to identify falls and locations of falls have been previously described and includes a robust approach to identifying overall injurious falls and to identifying falls for which syncope (heat-related and non-heat related syncope) and heat illness were contributing factors. (7) All EMS data entry into NEMSIS must abide by the standards set forth by the NHTSA Office of EMS and outlined in the NEMSIS data dictionary (https://nemsis.org/media/nemsis_v3/release-3.5.0/DataDictionary/PDFHTML/EMSDEMSTATE/index.html). Falls with EMS notations of seizures have been removed from the analyses.

### Variable coding

#### Sociodemographic data

The NEMSIS variables ePatient.13 and ePatient.15 were used to define patient sex (male, female) and age group (0-20, 21-50, 50-64 and 65+), respectively.

#### Alcohol and drug use data

The NEMSIS variable eHistory.17 contains data on the EMS clinician’s evaluation of whether alcohol or drugs were involved in an incident. Using these data patients were classified for the EMS clinician’s notation of evidence of: “alcohol use”, “drug use”, or “alcohol and drug use”. In the absence of a positive notation of substance use in the eHistory.17 variable, patients were classified as “evidence of substance use not noted”. The eHistory.17 variable includes multiple codes for each patient. Codes for the eHistory.17 variable of: 3117001, 3117005, 3117011, 3117009 were used to define “alcohol use”. Codes 3117003, 3117007, 3117009 were used to define “drug use”. If a patient had codes for both “alcohol use” and “drug use” they were coded as “alcohol and drug use”. NEMSIS codes for not applicable (7701001), not recorded (7701003), none reported (88010015), refused (8801019), and unable to complete (8801023) were used to create the “evidence of substance use not noted” category.

### Statistical analysis

Descriptive analyses for all EMS encounters for injurious falls by location and reported substance use were conducted, and data were cross tabulated by age group and sex. Syncope and heat illness are often causes of injurious falls and alcohol may play a different role in such falls than for falls that occurred without these co-morbidities, and falls without these comorbidities are more likely to be mechanical falls. (7, 37) As such, analyses were also conducted separately for injurious falls that included a notation of syncope, falls that included a notation of heat illness and falls that did not include notations of syncope or heat illness. We conducted all analyses in R Statistical Software (v4.3.1; R Core Team 2023).

### Results

In total 1,854,909 injuries from falls that required an EMS response were identified in the 2019 NEMSIS data. (7) Overall, for 7.4% (n=136,978, see table 1) of injurious falls there was a notation of substance use (alcohol and/or drug use). Notations of alcohol use alone made up 87.8% of the reports of substance use, with a notation of alcohol and drug use comprising 4.3% of reports involving substance use and drug use alone comprising 8.0% of these reports.

**Table 1.** Reported Alcohol and Drug Use Among Fall Patients by Location of Fall and by Presence of Syncope and Heat Illness.

| All Injurious Falls |  |  |  |  |  |  |
| --- | --- | --- | --- | --- | --- | --- |
| Substance Use | Overall<br>N = 1,854,909<sup>1</sup> | Indoors<br>N = 1,596,860<sup>1</sup> | Not Recorded<br>N = 51,355<sup>1</sup> | Indoor vs<br>Outdoor<br>Unclear,<br>N = 53,700<sup>1</sup> | Outdoors –<br>not street<br>N = 23,586<sup>1</sup> | Street or<br>Sidewalk<br>N = 129,408<sup>1</sup> |
| Alcohol | 120,262<br>(6.5%) | 86,950<br>(5.4%) | 1,494<br>(2.9%) | 6,023<br>(11.2%) | 1,857<br>(7.9%) | 23,938<br>(18.5%) |
| Alcohol and drugs | 5,823<br>(0.3%) | 4,160<br>(0.3%) | 106<br>(0.2%) | 245<br>(0.5%) | 107<br>(0.5%) | 1,205<br>(0.9%) |
| Drugs | 10,893<br>(0.6%) | 7,814<br>(0.5%) | 129<br>(0.3%) | 545<br>(1.0%) | 158<br>(0.7%) | 2,247<br>(1.7%) |
| Evidence of substance use not noted | 1,717,931<br>(92.6%) | 1,497,936<br>(93.8%) | 49,626<br>(96.6%) | 46,887<br>(87.3%) | 21,464<br>(91.0%) | 102,018<br>(78.8%) |
| Injurious falls with accompanying syncope |  |  |  |  |  |  |
|  | Overall<br>N = 73,600<sup>1</sup> | Indoors<br>N = 64,325<sup>1</sup> | Not Recorded<br>N = 1,402<sup>1</sup> | Indoor vs<br>Outdoor<br>Unclear<br>N = 2,351<sup>1</sup> | Outdoors -<br>not street<br>N = 800<sup>1</sup> | Street or<br>Sidewalk<br>N = 4,722<sup>1</sup> |
| Alcohol | 4,818<br>(6.5%) | 3,902<br>(6.1%) | 65<br>(4.6%) | 267<br>(11.4%) | 66<br>(8.3%) | 518<br>(11.0%) |
| Alcohol and drugs | 349<br>(0.5%) | 286<br>(0.4%) | 9<br>(0.6%) | 14<br>(0.6%) | 6<br>(0.8%) | 34<br>(0.7%) |
| Drugs | 916<br>(1.2%) | 755<br>(1.2%) | 12<br>(0.9%) | 49<br>(2.1%) | 6<br>(0.8%) | 94<br>(2.0%) |
| Evidence of substance use not noted | 67,517<br>(91.7%) | 59,382<br>(92.3%) | 1,316<br>(93.9%) | 2,021<br>(86.0%) | 722<br>(90.3%) | 4,076<br>(86.3%) |
| Injurious falls with accompanying heat illness |  |  |  |  |  |  |
|  | Overall<br>N = 9381 | Indoors<br>N = 6321 | Not Recorded<br>N = 171 | Indoor vs<br>Outdoor<br>Unclear<br>N = 421 | Outdoors -<br>not street<br>N = 541 | Street or<br>Sidewalk<br>N = 1931 |
| Alcohol | 55<br>(5.9%) | 30<br>(4.7%) | 2<br>(11.8%) | 5<br>(11.9%) | 3<br>(5.6%) | 15<br>(7.8%) |
| Alcohol<br>and drugs | 4<br>(0.4%) | 3<br>(0.5%) | 0<br>(0.0%) | 1<br>(2.4%) | 0<br>(0.0%) | 0<br>(0.0%) |
| Drugs | 13<br>(1.4%) | 6<br>(0.9%) | 0<br>(0.0%) | 1<br>(2.4%) | 1<br>(1.9%) | 5<br>(2.6%) |
| Evidence<br>of<br>substance<br>use not<br>noted | 866<br>(92.3%) | 593<br>(93.8%) | 15<br>(88.2%) | 35<br>(83.3%) | 50<br>(92.6%) | 173<br>(89.6%) |
| Injurious falls without accompanying syncope or heat illness |  |  |  |  |  |  |
|  | **Overall**<br>**N = 1,780,371<sup>1</sup>** | **Indoors**<br>**N =**<br>**1,531,903<sup>1</sup>** | **Not Recorded**<br>**N = 49,936<sup>1</sup>** | **Indoor vs**<br>**Outdoor**<br>**Unclear**<br>**N = 51,307<sup>1</sup>** | **Outdoors -**<br>**not street**<br>**N = 22,732<sup>1</sup>** | **Street or**<br>**Sidewalk**<br>**N = 124,493<sup>1</sup>** |
| Alcohol | 115,389<br>(6.5%) | 83,018<br>(5.4%) | 1,427<br>(2.9%) | 5,751<br>(11.2%) | 1,788<br>(7.9%) | 23,405<br>(18.8%) |
| Alcohol<br>and drugs | 5,470<br>(0.3%) | 3,871<br>(0.3%) | 97<br>(0.2%) | 230<br>(0.4%) | 101<br>(0.4%) | 1,171<br>(0.9%) |
| Drugs | 9,964<br>(0.6%) | 7,053<br>(0.5%) | 117<br>(0.2%) | 495<br>(1.0%) | 151<br>(0.7%) | 2,148<br>(1.7%) |
| Evidence<br>of<br>substance<br>use not<br>noted | 1,649,548<br>(92.7%) | 1,437,961<br>(93.9%) | 48,295<br>(96.7%) | 44,831<br>(87.4%) | 20,692<br>(91.0%) | 97,769<br>(78.5%) |

<sup>1</sup>n (%)

For falls that occurred on a street or sidewalk, there was a notation of substance use for 21.2% (n= 27,390) of patients, with alcohol use alone making up 87.4% (n=23,938) of these reports. When injurious falls were separated into falls with a notation of syncope, falls with a notation of heat illness and falls without notation of syncope or heat illness, the overall prevalences of alcohol and/or drug involvement was similar across the three groups (see Table 1). However, for falls occurring on streets and sidewalks, the prevalence of alcohol involvement alone, at 18.8%, was highest for injurious falls without accompanying syncope or heat illness. Across almost all age groups, for both fall types, the prevalence of alcohol involvement was higher for falls occurring on streets and sidewalks compared to falls occurring indoors. For falls with accompanying syncope, that occurred on streets and sidewalks, adults ages 21-50 and 51-64 had the highest proportion of falls reported to involve alcohol use alone (12.3% and 18.1% respectively). Among those experiencing falls on streets and sidewalks without accompanying syncope or heat illness, these age groups also had the highest proportion of falls reported to involve alcohol (25.3% and 27.7% respectively).

Table 2 shows age stratified analyses for falls with a notation of accompanying syncope and Table 3 shows age stratified analyses for falls without accompanying syncope or heat illness (there were not enough falls with a notation of heat illness for meaningful age stratification).

**Table 2.**
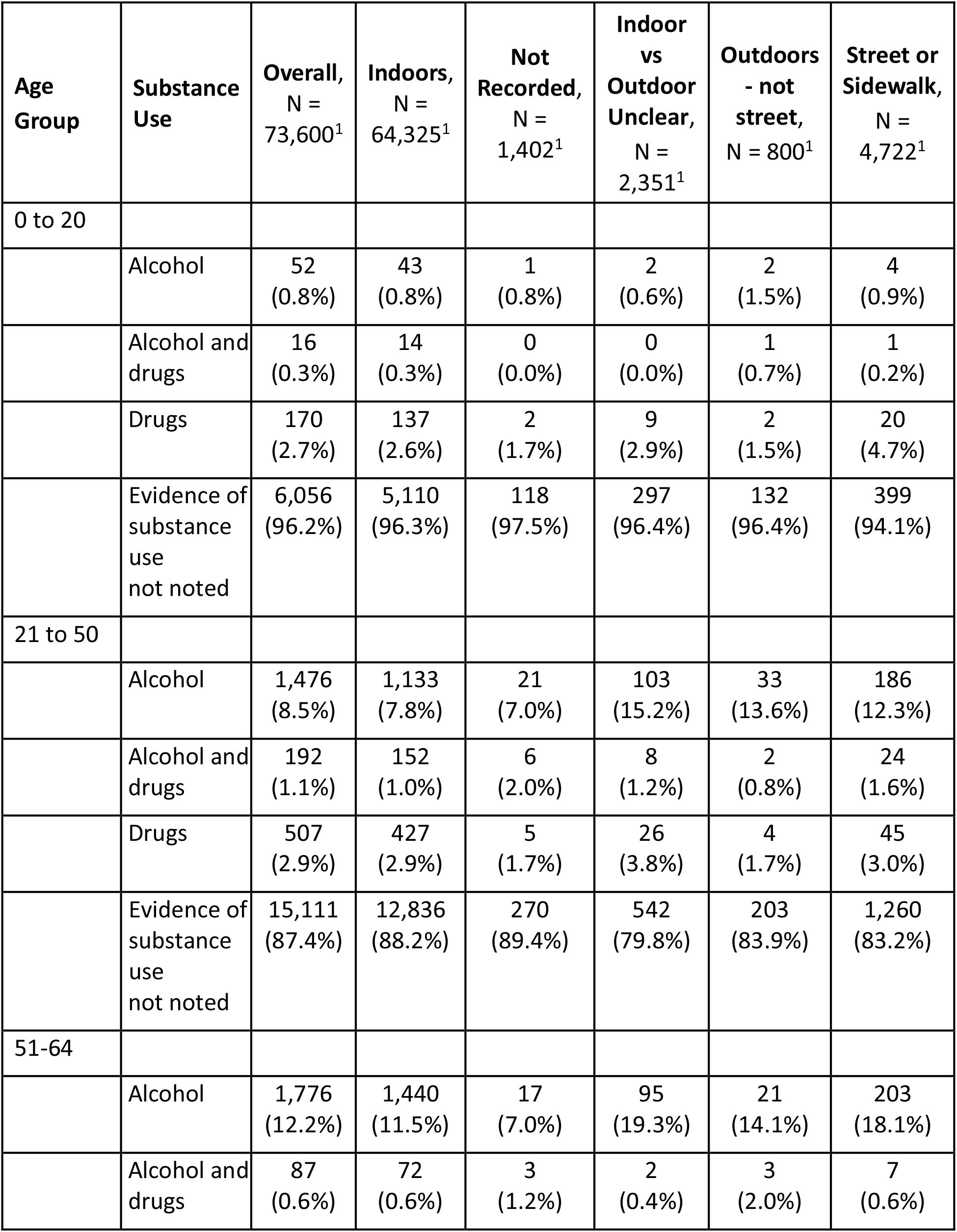
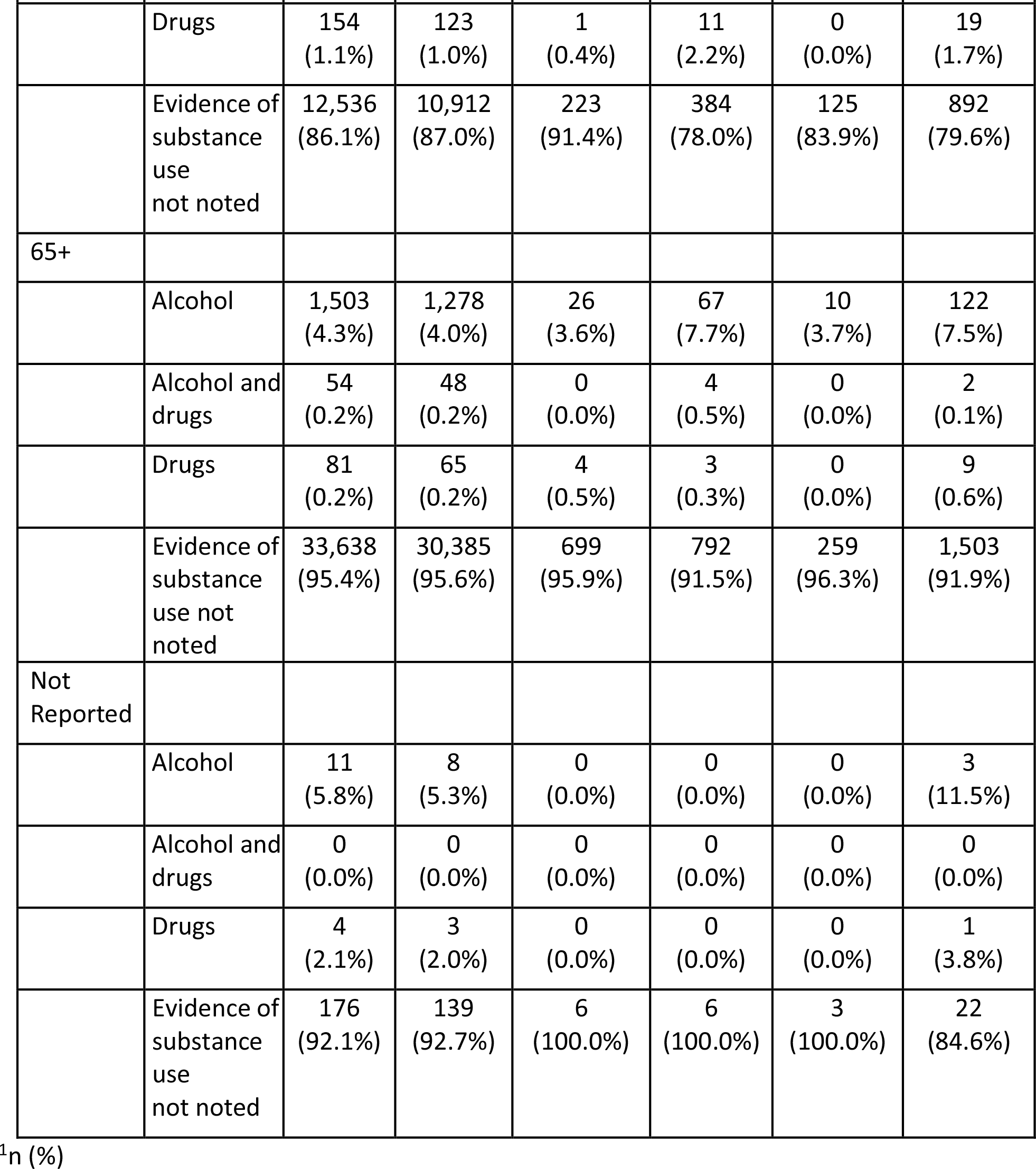
Substance use by Patient Age and Location, among Falls Patients with Accompanying Syncope.

**Table 3.** Substance use by Patient Age and Location, among Falls Patients without Accompanying Syncope or Heat-Illness.

| **Age Group** | **Substance Use** | **Overall,**<br>**N =**<br>**1,780,371<sup>1</sup>** | **Indoors,**<br>**N =**<br>**1,531,903<sup>1</sup>** | **Not Recorded**<br>**, N =**<br>**49,936<sup>1</sup>** | **Indoor**<br>**vs**<br>**Outdoor**<br>**Unclear**<br>**,**<br>**N =**<br>**51,307<sup>1</sup>** | **Outdoors - not**<br>**street,**<br>**N =**<br>**22,732<sup>1</sup>** | **Street or**<br>**Sidewalk**<br>**,**<br>**N =**<br>**124,493<sup>1</sup>** |
| --- | --- | --- | --- | --- | --- | --- | --- |
| <21 |  |  |  |  |  |  |  |
|  | Alcohol | 2,039<br>(2.0%) | 1,332<br>(1.7%) | 26<br>(1.0%) | 130<br>(1.5%) | 74<br>(1.1%) | 477<br>(5.8%) |
|  | Alcohol and drugs | 264<br>(0.3%) | 178<br>(0.2%) | 8<br>(0.3%) | 15<br>(0.2%) | 16<br>(0.2%) | 47<br>(0.6%) |
|  | Drugs | 718<br>(0.7%) | 486<br>(0.6%) | 18<br>(0.7%) | 54<br>(0.6%) | 21<br>(0.3%) | 139<br>(1.7%) |
|  | Evidence of substance use not noted | 100,948<br>(97.1%) | 76,054<br>(97.4%) | 2,518<br>(98.0%) | 8,327<br>(97.7%) | 6,493<br>(98.3%) | 7,556<br>(91.9%) |
| 21 – 50 |  |  |  |  |  |  |  |
|  | Alcohol | 38,150<br>(15.9%) | 24,484<br>(13.9%) | 471<br>(8.2%) | 2,455<br>(18.8%) | 800<br>(13.4%) | 9,940<br>(25.3%) |
|  | Alcohol and drugs | 2,333<br>(1.0%) | 1,449<br>(0.8%) | 44<br>(0.8%) | 130<br>(1.0%) | 52<br>(0.9%) | 658<br>(1.7%) |
|  | Drugs | 5,444<br>(2.3%) | 3,469<br>(2.0%) | 58<br>(1.0%) | 279<br>(2.1%) | 96<br>(1.6%) | 1,542<br>(3.9%) |
|  | Evidence of substance use not noted | 193,777<br>(80.8%) | 146,161<br>(83.3%) | 5,157<br>(90.0%) | 10,216<br>(78.1%) | 5,027<br>(84.1%) | 27,216<br>(69.2%) |
| 51-64 |  |  |  |  |  |  |  |
|  | Alcohol | 42,537<br>(14.4%) | 30,207<br>(12.6%) | 467<br>(6.8%) | 2,068<br>(19.7%) | 590<br>(14.6%) | 9,205<br>(27.7%) |
|  | Alcohol<br>and<br>drugs | 1,562<br>(0.5%) | 1,100<br>(0.5%) | 19<br>(0.3%) | 63<br>(0.6%) | 26<br>(0.6%) | 354<br>(1.1%) |
|  | Drugs | 2,094<br>(0.7%) | 1,556<br>(0.6%) | 22<br>(0.3%) | 128<br>(1.2%) | 28<br>(0.7%) | 360<br>(1.1%) |
|  | Evidence<br>of<br>substance<br>use<br>not noted | 248,822<br>(84.3%) | 207,519<br>(86.3%) | 6,347<br>(92.6%) | 8,233<br>(78.5%) | 3,393<br>(84.0%) | 23,330<br>(70.2%) |
| 65+ |  |  |  |  |  |  |  |
|  | Alcohol | 32,120<br>(2.8%) | 26,653<br>(2.6%) | 454<br>(1.3%) | 1,083<br>(5.7%) | 313<br>(5.3%) | 3,617<br>(8.5%) |
|  | Alcohol<br>and<br>drugs | 1,279<br>(0.1%) | 1,121<br>(0.1%) | 26<br>(0.1%) | 22<br>(0.1%) | 6<br>(0.1%) | 104<br>(0.2%) |
|  | Drugs | 1,656<br>(0.1%) | 1,507<br>(0.1%) | 18<br>(0.1%) | 34<br>(0.2%) | 5<br>(0.1%) | 92<br>(0.2%) |
|  | Evidence<br>of<br>substance<br>use<br>not<br>noted | 1,093,503<br>(96.9%) | 997,505<br>(97.1%) | 33,826<br>(98.5%) | 17,742<br>(94.0%) | 5,627<br>(94.6%) | 38,803<br>(91.1%) |
| Not<br>Reported |  |  |  |  |  |  |  |
|  | Alcohol | 543<br>(4.1%) | 342<br>(3.1%) | 9<br>(2.0%) | 15<br>(4.6%) | 11<br>(6.7%) | 166<br>(15.8%) |
|  | Alcohol<br>and<br>drugs | 32<br>(0.2%) | 23<br>(0.2%) | 0<br>(0.0%) | 0<br>(0.0%) | 1<br>(0.6%) | 8<br>(0.8%) |
|  | Drugs | 52<br>(0.4%) | 35<br>(0.3%) | 1<br>(0.2%) | 0<br>(0.0%) | 1<br>(0.6%) | 15<br>(1.4%) |
|  | Evidence<br>of<br>substance<br>use<br>not noted | 12,498<br>(95.2%) | 10,722<br>(96.4%) | 447<br>(97.8%) | 313<br>(95.4%) | 152<br>(92.1%) | 864<br>(82.1%) |

<sup>1</sup>n (%)

For falls with accompanying syncope and falls without accompanying syncope or heat illness, regardless of location, EMS reports for male patients were more likely to include a report of alcohol and/or drugs than for female patients (see Table 3 and 4). Also for both fall types, the highest prevalences of substance use were found among men for falls occurring on streets and sidewalks. The highest prevalence of alcohol use alone, at 25.1%, was found among men experiencing falls on streets and sidewalks, without accompanying syncope or heat illness.

**Table 4.** Substance use by Patient Sex and Location, among Falls Patients with Accompanying Syncope.

| **Patient Sex** | **Substance Use** | **Overall**<br>**N =**<br>**73,600<sup>1</sup>** | **Indoors**<br>**N =**<br>**64,325<sup>1</sup>** | **Not**<br>**Recorded**<br>**N =**<br>**1,402<sup>1</sup>** | **Indoor vs**<br>**Outdoor**<br>**Unclear**<br>**N =**<br>**2,351<sup>1</sup>** | **Outdoors**<br>**- not**<br>**street**<br>**N = 800<sup>1</sup>** | **Street or**<br>**Sidewalk**<br>**N =**<br>**4,722<sup>1</sup>** |
| --- | --- | --- | --- | --- | --- | --- | --- |
| Female |  |  |  |  |  |  |  |
|  | Alcohol | 1,519<br>(4.1%) | 1,260<br>(3.8%) | 21<br>(3.1%) | 86<br>(8.3%) | 17<br>(4.5%) | 135<br>(6.5%) |
|  | Alcohol<br>and drugs | 115<br>(0.3%) | 90<br>(0.3%) | 2<br>(0.3%) | 6<br>(0.6%) | 4<br>(1.0%) | 13<br>(0.6%) |
|  | Drugs | 361<br>(1.0%) | 310<br>(0.9%) | 2<br>(0.3%) | 15<br>(1.4%) | 5<br>(1.3%) | 29<br>(1.4%) |
|  | Evidence<br>of<br>substance<br>use not<br>noted | 34,959<br>(94.6%) | 31,125<br>(94.9%) | 648<br>(96.3%) | 931<br>(89.7%) | 355<br>(93.2%) | 1,900<br>(91.5%) |
| Male |  |  |  |  |  |  |  |
|  | Alcohol | 3,286<br>(9.0%) | 2,633<br>(8.4%) | 44<br>(6.1%) | 180<br>(13.8%) | 48<br>(11.6%) | 381<br>(14.5%) |
|  | Alcohol<br>and drugs | 234<br>(0.6%) | 196<br>(0.6%) | 7<br>(1.0%) | 8<br>(0.6%) | 2<br>(0.5%) | 21<br>(0.8%) |
|  | Drugs | 553<br>(1.5%) | 443<br>(1.4%) | 10<br>(1.4%) | 34<br>(2.6%) | 1<br>(0.2%) | 65<br>(2.5%) |
|  | Evidence<br>of<br>substance<br>use not<br>noted | 32,355<br>(88.8%) | 28,081<br>(89.6%) | 661<br>(91.6%) | 1,083<br>(83.0%) | 363<br>(87.7%) | 2,167<br>(82.3%) |
| Not<br>Reported |  |  |  |  |  |  |  |
|  | Alcohol | 13<br>(6.0%) | 9<br>(4.8%) | 0<br>(0.0%) | 1<br>(12.5%) | 1<br>(20.0%) | 2<br>(18.2%) |
|  | Alcohol<br>and drugs | 0<br>(0.0%) | 0<br>(0.0%) | 0<br>(0.0%) | 0<br>(0.0%) | 0<br>(0.0%) | 0<br>(0.0%) |
|  | Drugs | 2<br>(0.9%) | 2<br>(1.1%) | 0<br>(0.0%) | 0<br>(0.0%) | 0<br>(0.0%) | 0<br>(0.0%) |
|  | Evidence<br>of<br>substance<br>use not<br>noted | 203<br>(93.1%) | 176<br>(94.1%) | 7<br>(100.0%) | 7<br>(87.5%) | 4<br>(80.0%) | 9<br>(81.8%) |

<sup>1</sup>n (%)

**Table 5.** Substance use by Patient Sex and Location, among Falls Patients without Accompanying Syncope or Heat-Illness.

| Sex | Substance Use | Overall<br>N =<br>1,780,371<sup>1</sup> | Indoors<br>N =<br>1,531,903<sup>1</sup> | Not<br>Recorded<br>N =<br>49,936<sup>1</sup> | Indoor<br>vs<br>Outdoor<br>Unclear<br>N =<br>51,307<sup>1</sup> | Outdoors - not<br>street<br>N =<br>22,732<sup>1</sup> | Street or<br>Sidewalk<br>N =<br>124,493<sup>1</sup> |
| --- | --- | --- | --- | --- | --- | --- | --- |
| Female |  |  |  |  |  |  |  |
|  | Alcohol | 38,742<br>(3.8%) | 30,677<br>(3.4%) | 511<br>(1.8%) | 1,761<br>(6.9%) | 554<br>(5.2%) | 5,239<br>(10.1%) |
|  | Alcohol<br>and drugs | 1,915<br>(0.2%) | 1,543<br>(0.2%) | 33<br>(0.1%) | 59<br>(0.2%) | 26<br>(0.2%) | 254<br>(0.5%) |
|  | Drugs | 3,589<br>(0.3%) | 2,872<br>(0.3%) | 46<br>(0.2%) | 131<br>(0.5%) | 38<br>(0.4%) | 502<br>(1.0%) |
|  | Evidence of<br>substance<br>use not<br>noted | 985,895<br>(95.7%) | 878,008<br>(96.2%) | 28,371<br>(98.0%) | 23,524<br>(92.3%) | 10,047<br>(94.2%) | 45,945<br>(88.5%) |
| Male |  |  |  |  |  |  |  |
|  | Alcohol | 76,253<br>(10.3%) | 52,051<br>(8.5%) | 911<br>(4.4%) | 3,973<br>(15.5%) | 1,228<br>(10.3%) | 18,090<br>(25.1%) |
|  | Alcohol<br>and drugs | 3,543<br>(0.5%) | 2,321<br>(0.4%) | 64<br>(0.3%) | 171<br>(0.7%) | 74<br>(0.6%) | 913<br>(1.3%) |
|  | Drugs | 6,329<br>(0.9%) | 4,153<br>(0.7%) | 71<br>(0.3%) | 364<br>(1.4%) | 111<br>(0.9%) | 1,630<br>(2.3%) |
|  | Evidence of<br>substance<br>use not<br>noted | 656,868<br>(88.4%) | 554,457<br>(90.5%) | 19,442<br>(94.9%) | 21,107<br>(82.4%) | 10,553<br>(88.2%) | 51,309<br>(71.3%) |
| Not<br>Reported |  |  |  |  |  |  |  |
|  | Alcohol | 394<br>(5.4%) | 290<br>(5.0%) | 5<br>(1.0%) | 17<br>(7.8%) | 6<br>(5.9%) | 76<br>(12.4%) |
|  | Alcohol<br>and drugs | 12<br>(0.2%) | 7<br>(0.1%) | 0<br>(0.0%) | 0<br>(0.0%) | 1<br>(1.0%) | 4<br>(0.7%) |
|  | Drugs | 46<br>(0.6%) | 28<br>(0.5%) | 0<br>(0.0%) | 0<br>(0.0%) | 2<br>(2.0%) | 16<br>(2.6%) |
|  | Evidence of<br>substance<br>use not<br>noted | 6,785<br>(93.8%) | 5,496<br>(94.4%) | 482<br>(99.0%) | 200<br>(92.2%) | 92<br>(91.1%) | 515<br>(84.3%) |

<sup>1</sup>n (%)

### Discussion

Overall, 7.4% of all falls requiring EMS response in 2019 involved reported alcohol and/or drug use, while 21.2% of injurious falls occurring on streets or sidewalks involved reported substance use. For all fall types, alcohol use alone was the most frequently reported substance involved in the fall. The prevalence of alcohol involvement for falls that occurred on streets and sidewalks was highest among falls patients who did not have accompanying syncope or heat illness. Falls without an accompanying notation of syncope or heat illness, among adults ages 21-64 and among men were more likely to reportedly involve alcohol alone compared to older adults or youth and women. Falls that occurred on streets or sidewalks were consistently more likely to reportedly involve alcohol across age categories and sexes compared to falls than occurred indoors.

The estimates of the overall prevalence of alcohol and substance use among falls patients presented here are consistent with recent reports using other datasets. Analyses of the 2019– 2020 National Hospital Ambulatory Medical Care Survey (NHAMCS) found that 8.4% of fall-related ED visits had indications of alcohol and/or substance use. (38) Analyses of the 2011 to 2020 National Electronic Injury Surveillance System-All Injury Program (NEISS) for patients seeking treatment at EDs found that alcohol involved falls comprised 2.2% of ED visits for falls among patients age 65 and older. (1) This prevalence is consistent with the overall proportion of alcohol involved falls in this age group observed in the NEMSIS data (2.8% for falls without syncope or heat illness). Like prior studies, the analyses of the NHAMCS and NEISS data did not sub-divide falls by location of occurrence and did not separately analyze falls by indications of accompanying syncope and heat illness. (1) Fall injuries that do not have these co-morbidities are more likely to be mechanical falls, for which alcohol consumption, and the resultant loss in coordination, may play a larger role. (37)

These findings suggest that alcohol use is present in a large proportion of falls that occur on streets and sidewalks, particularly among men and adults between the ages of 21 and 64. Falls that occur on streets and sidewalks that involve alcohol may be in close proximity to businesses that serve alcohol. This is consistent with previous findings that suggest a higher density of alcohol serving establishments is associated with both increased alcohol consumption (39) and increased pedestrian injury from motor vehicles. (14, 40-42) Further, the increased prevalence of men and adults ages 21-64 among injurious falls involving alcohol is consistent with the higher prevalence of alcohol consumption in these groups in the US. (14, 43) There are several possible policy options that could be used to address the burden of injurious falls occurring on streets and sidewalks that involve alcohol. (12) Such options include considering appropriate lighting for commercial areas, improved maintenance of sidewalks and curb cuts and improved maintenance of roadbeds at street crossings. (44) States and cities might also re-evaluate policies around nightlife districts and open container laws that encourage the consumption of alcohol while walking. (12, 20, 26) Lastly, it is possible that individuals experiencing homelessness are at risk for experiencing alcohol involved injurious falls in public spaces. (45) If indeed individuals experiencing homelessness do comprise a large proportion of these falls patients, the medical costs of such falls should be factored in to debates on funding homelessness prevention programs.

This is the first study to describe the burden of substance use involved falls occurring on streets and sidewalks from a well-documented national sample. These data are a first step at informing public policies and design strategies aimed at preventing alcohol-involved falls. However, there are also limitations that should be noted. First, we labelled all EMS reports where alcohol or drug use were not noted as “evidence of substance use not noted”, and functionally in our calculations of proportions treated them as indicating an absence of substance use. However, it is unlikely that EMS clinicians’ notations of substance use based on patient’s on-scene reports or physical evidence at the scene is a highly sensitive measure of substance use by patients. Therefore, these positive reports of substance involvement in NEMSIS are likely to be an underestimate of the true prevalence of the involvement of substance use among patients treated for injurious falls. Furthermore, there are likely a large number of additional injurious falls involving alcohol and drugs, where the individuals either do not seek EMS care, perhaps due to stigma surrounding alcohol overconsumption and drug use, or those who seek care directly from a hospital or urgent care facility. Taken together, these limitations likely resulted in an underestimation of the burden of substance use involved pedestrian falls.

In conclusion, this study suggests that a large proportion of falls that occur outdoors on streets and sidewalks and require EMS response involve substance use. Research is needed to assess the link between venues serving alcohol, the built environments around these venues and risk of pedestrian falls and to examine if policies to create nightlife districts lead to an increase in pedestrian falls.

### Authors’ contributions

Nicole G. Itzkowitz – conducted data analyses and wrote initial drafts of the manuscript. Kathryn G. Burford – conducted data analyses and edited and revised the manuscript.

Remle P Crowe – advised on defining falls in NEMSIS, conceptualized the study and wrote and edited portions of the manuscript.

Henry E Wang – advised on defining falls in NEMSIS and on data analyses and edited the manuscript.

Alexander X Lo – advised on defining syncope, heat illness and seizures in the NEMSIS data, helped direct the data analyses and edited the manuscript.

Andrew G. Rundle – conceptualized the study, conducted data analyses to identify patients, advised on data analysis and edited the manuscript.

## Data Availability

All data produced are available online at https://nemsis.org/using-ems-data/request-research-data/

## Acknowledgements

National Highway Traffic Safety Administration (NHTSA), National Emergency Medical Services Information System (NEMSIS). The content reproduced from the NEMSIS Database remains the property of the National Highway Traffic Safety Administration (NHTSA). The National Highway Traffic Safety Administration is not responsible for any claims arising from works based on the original Data, Text, Tables, or Figures.

